# From Reductionist Skills to Meaningful Learning: Trust and Humility in Bedside Cardiac Assessment

**DOI:** 10.1101/2022.08.23.22279141

**Authors:** James L. Meisel, Deborah D. Navedo, Isaac O. Opole, Gail March Cohen, Sheilah A. Bernard, Hugo Carmona, Ahmed H. Nahas, Carly M. Eiduson, Nick Papps

## Abstract

**PURPOSE:** Notions of trust are foundational to competency-based medical education. “Entrustability” underlies assessment; assessment is guided by integration into curricula of learners’ knowledge, skills, and attitudes. However, attitudinal notions of trust are not commonly conceptualized as integral to such frameworks. Overlap between concepts of entrustability and trust as an attitude creates opportunity to infuse trust into competency frameworks.

We explored how an original bedside cardiac assessment (BCA) curriculum that supported professional attitudes, knowledge, and skills scaffolded clinical learning in a cohort of internal medicine clerkship students.

**METHODS:** The curriculum urged students to hear patients’ perspectives with humility and as key to diagnostic reasoning. Assigned short videos preceded two facilitated classes that included discussing a patient’s startling question, “Why should I trust your clinical skills?” and recognizing, in simulated clinical encounters, disparate patients’ perspectives.

To better understand their experiences, we asked sixty-seven students to complete two post-class open-ended questions. We analyzed responses using content and thematic analyses.

**RESULTS:** Emergent codes clustered around themes in two categories: “Successful Learning” around effective learning strategies and meaningful peer encounters, skills practice, and educator encounters; and “Opportunities for Improvement,” around instructional design, learning preferences, and instruction-related improvements.

**CONCLUSION:** Themes suggested effective learning and meaningful interactions. Comments affirmed the importance of attitudinal aspects of skills development; human interaction while learning; and humility, a linchpin of expertise development and patient-centered communication. All contribute to professional identity formation (PIF). Instructional design improvements were incorporated into the final version of the curriculum. Limitations included inability to examine nuances of emergent themes from the limited data set.

We are studying the curriculum’s effects on BCA-related knowledge, skills, and attitudes and trust-worthiness as a learning construct. Research opportunities include impacts on humility, patient-centeredness, and PIF. We hope this exploratory work will stimulate conversations around expanded roles of notions of trust in medical education.

**Plain-language Summary:** *Why was the study done?:* The patient–doctor relationship is built upon trust in the doctor’s knowledge, skills, and attitudes. In this study, researchers explored medical students’ experience of participating in a curriculum that encouraged them to explore such attitudes as humility and trust (“Why should I trust your clinical skills?”) while learning to care for patients with heart problems.

*What did the researchers do and find?:* The researchers used an educational approach in which the process of learning is seen as meaningful, not as simply learning isolated facts or skills. To better understand students’ experience of participating in an early version of the curriculum, researchers asked sixty-seven students to complete open-ended questions after two sets of class activities. They then analyzed students’ responses, looking for recurring themes. The analysis suggested that the learning strategies were effective and that learning with peers; skills practice; and interacting with educators were meaningful. The analysis also revealed several opportunities to improve the curriculum itself.

*What do these results mean?:* Students reported that human interaction meaningfully contributed to successful learning. In particular, humility was key to patient-centered communication and building trust. All of this suggests the curriculum may help students develop identities as trustworthy professionals. Given a relatively small dataset, a limitation of this paper was that the researchers could not explore further nuance. The researchers anticipate that this work will stimulate conversations around expanded roles of trust in medical education.

## Introduction

Notions of trust are foundational to the concept of competency-based medical education (CBME).^1^ “Entrustability” is typically discussed in the context of assessment; assessment is guided by integration into curricula of learners’ knowledge, skills, and attitudes.^2,3^ However, attitudes, which complement knowledge and skill acquisition, are not commonly conceptualized as notions of trust and humility that are integral to competency frameworks.^4–8^ Overlap between the concepts of trust as an attitude and entrustability creates an opportunity to meaningfully infuse trust into competency development frameworks (Figure 1).

**Figure 1:** Notions of trust as a central element of both competencies and entrustment decision-making in competency-based medical education Abbreviation: EPAs, entrustable professional activities

Bedside cardiac assessment (BCA), which requires the integration of patients’ physiology, symptoms, signs, and social and psychological aspects of illness, exemplifies one such set of knowledge, skills, and attitudes. When performed well, BCA also facilitates faster and more expedient diagnosis and improves the performance of diagnostic technology like focused cardiac ultrasound. However, teaching and learning BCA are declining.^9^ Traditional, reductionist approaches address knowledge of cardiac pathophysiology and skills such as auscultation, jugular venous pressure measurement, or point-of-care ultrasonography.^9^ Competent BCA also requires integration of diagnostic reasoning skills and self-aware attitudes such as the importance of humility, patient-centered communication, and trust from the patient’s perspective.^8,10,11^

CBME curricula encompass a spectrum of relationships – between learners and supervisors, trainees and their programs, and programs and society,^12^ but current constructs may not adequately address or enhance the meaning that individual learners make of their perceived trust-worthiness to patients. In a commentary entitled, “Entrustable Professional Activities - Not Just for Assessment,” Turner broaches this, asking what support and resources learners need to further their development, while noting that individual learning plans should arise from curricula based on desired patient outcomes.^3^ Karp et al explored clerkship students’ perception of trust in them from supervisors, not patients. Notably, they identified themes related to the importance of trust as scaffolding for learning; on the learning environment; and on perceived consequences for patients.^13^ Both support the guiding tenet of the Coalition for Physician Accountability’s Undergraduate Medical Education-Graduate Medical Education [UME-GME] Review Committee that “[A]bove all else, [principles supporting] the UME-GME transition must optimally serve the public good.”^1^ In an editorial accompanying Karp’s exploration, “Trust as a Scaffold for Competency-Based Medical Education,” Young and Elnicki observe, “…when we conceptualize CBME through the lens of trusting relationships, greater layers of nuance and value begin to emerge.”^12^ Notions of trust counter the “check-box approach of CBME that has been said to reduce the medical profession to a series of superficial skills,”^5^ with physician humility, in particular, fundamental to building patient trust.^8^ We conceptualized students’ attitudes about trust-worthiness as important to learning a more holistic^10^ and effective approach to BCA. The purpose of this study was to explore the meaning that a cohort of internal medicine clerkship students would make from their experience of participating in a BCA competency development framework into which notions of trust, opportunities for attitudinal (affective)^5^ growth, and a holistic approach to patient care^14^ had been incorporated.

## Methods

### Development of the curriculum

We deliberated what it meant to improve teaching and learning BCA as a fully developed competency, ultimately focusing on learning outcomes to answer a patient’s question, “Why should I trust your clinical skills?” To develop the curriculum, we used constructivist learning theories to support acquisition of professional attitudes as well as traditional BCA knowledge and skills like diagnostic reasoning and auscultation.^9^ Constructivism is a cognitive learning theory in which learning is not simply “knowledge to be attained,” but rather a meaning-making process based upon the learner’s prior knowledge and experience or opportunities to engage with others about shared problems.^11^ We sought to help learners go beyond facts to gain deep understanding that could then be applied in new contexts.^15^

Explicitly incorporating notions of trust, the curriculum and its title, *Listen Before You Auscultate*,^16^ urge learners to hear the patient’s perspective with humility, as a human predicament,^8^ and diagnostically, as an element of the clinical history in diagnostic reasoning. The development of both are central to the progression of expertise,^8^ so we designed the assignments and activities to enable participants to learn and practice tools such as hypothesis testing and pattern recognition in various contexts.

We used several, more fine-tuned instructional design approaches to foster the development of professional growth, including multiple opportunities for learners to self-assess their knowledge, beliefs,^8^ and diagnostic reasoning abilities. With conceptual change theory, for example, we sought to generate dissatisfaction with learners’ current conceptions and skills, ensure that new material would be both understandable and plausible, and create the expectation that the time invested would be fruitful.^17^ An emphasis on understanding causal relationships, another hallmark of the expertise development^18^ that is integral to CBME^2^ and professional growth, further helps learners process patterns more efficiently.

In the flipped class curriculum, clerkship directors assigned pre-class, formative self-assessments and short videos followed by two, one-hour sets of in-class activities (Figure 2). In the first video, a patient unexpectedly asked the viewer, “Why should I trust your clinical skills?” followed by an onscreen instructor emphasizing the role of empathy in patient-centered communication. The facilitated class sessions included a large-group discussion about the patient’s startling question and simulated clinical encounters. Scenarios in the latter differed only by the perspective that the patient - a scared builder, an anxious biochemistry professor, or a kind patient from Latin America - brought to them. The activity gave students opportunities to demonstrate expertise and “[admit] limitations in social interactions”; both are challenges to cultivating physician humility.^8^ Encounters were most successful when students elicited their patients’ perspectives.

**Figure 2:** *Listen Before You Auscultate* bedside cardiac assessment curriculum Abbreviation: BCA, bedside cardiac assessment Meisel JL, Chen DCR, Cohen GM, et al. Appendix C: Curriculum overview. In: Listen before you auscultate: an active-learning approach to bedside cardiac assessment. *MedEdPORTAL*. 2023;19:11362 © 2023 Meisel JL, Chen DCR, Cohen GM, et al. Distributed under the terms of the Creative Commons CC BY-NC 4.0 license

### Evaluation strategy

In the first of a two-year pilot, we implemented the integrated curriculum at Boston University Medical Center (BUMC, Massachusetts USA), Kansas University Medical Center (Kansas USA), Central Clinical School Monash University (Australia), and Tan Tock Seng Hospital (Singapore). To explore learners’ experiences, we surveyed the third-year students on internal medicine rotation at BUMC and analyzed their short-answer responses using qualitative methodologies.

From November 2017 to June 2018, we asked 67 third-year medicine students to complete post-class surveys that included open-ended questions. After the first classroom session, we surveyed students about their experiences with the pre-class assignments and the session. After the second session, we surveyed students about their experiences in that session and the curriculum as a whole.

Minor changes to the survey questions resulted in responses that were more actionable for continuous quality improvement of the curriculum. Because the open-ended questions varied slightly, content analysis^19^ allowed us to group these questions into two categories, those related to successful learning and those related to opportunities for improvement (Table 1).

**Table 1.**
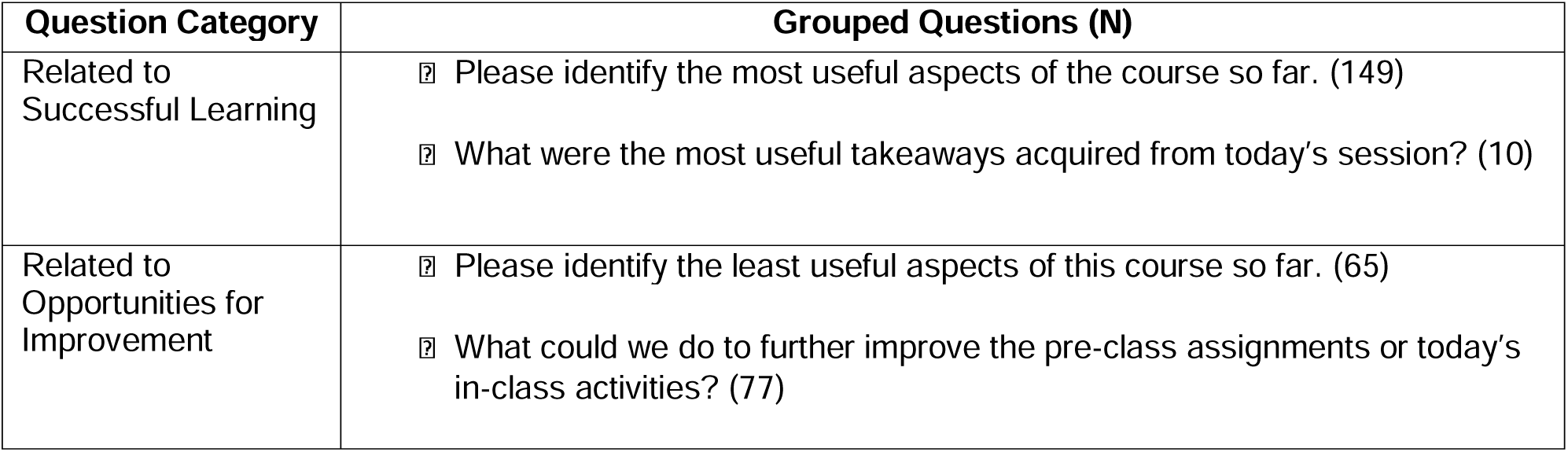
Question Categories that Resulted from the Grouped Survey Questions, This analysis and categorization was required due to variation in the survey questions presented to the students following the activity. See text for details.

We then analyzed the responses using a sequence of content analysis and then thematic analysis.^20^ Applied Thematic Analysis^21^ approach was implemented in which two researchers (DN, IO) independently engaged the data using an initial open coding strategy. Codes were developed using a constant comparison methodology, and we developed the final codebook based on consensus discussions to resolve differences. A third researcher (JM) served as arbiter to facilitate differences in perspectives. A re-examination of the codes in context resulted in the emergence of themes, reported below.

## Results

At the end of each of the two in-person classes, each participant was offered a survey that included the two open-ended responses. This yielded a total of 489 open-ended responses, of which 61.6% (n=301) were analyzed. The remaining 38.4% (n=188) were blank or unintelligible. There was a 75.4% (n=159/211) completion rate for questions related to “Successful Learning” and a 51.1% (n=142/278) completion rate for questions related to “Opportunities for Improvement.” First-round coding agreement rate was 54.8% (n=165/301). All coding disagreements were resolved by discussion resulting in consensus, except for 3.7% (n=11/301), which required arbitration. An interrater reliability analysis was performed between the final coding of the two raters. For this purpose, we used Cohen’s kappa, a measure of agreement in projects with categorical variables. Cohen’s kappa was 0.91, showing very strong final agreement in the final round of coding. Thus, common definitions of codes emerged easily, resulting in effective codebooks (Tables 2 and 3).

**Table 2.**
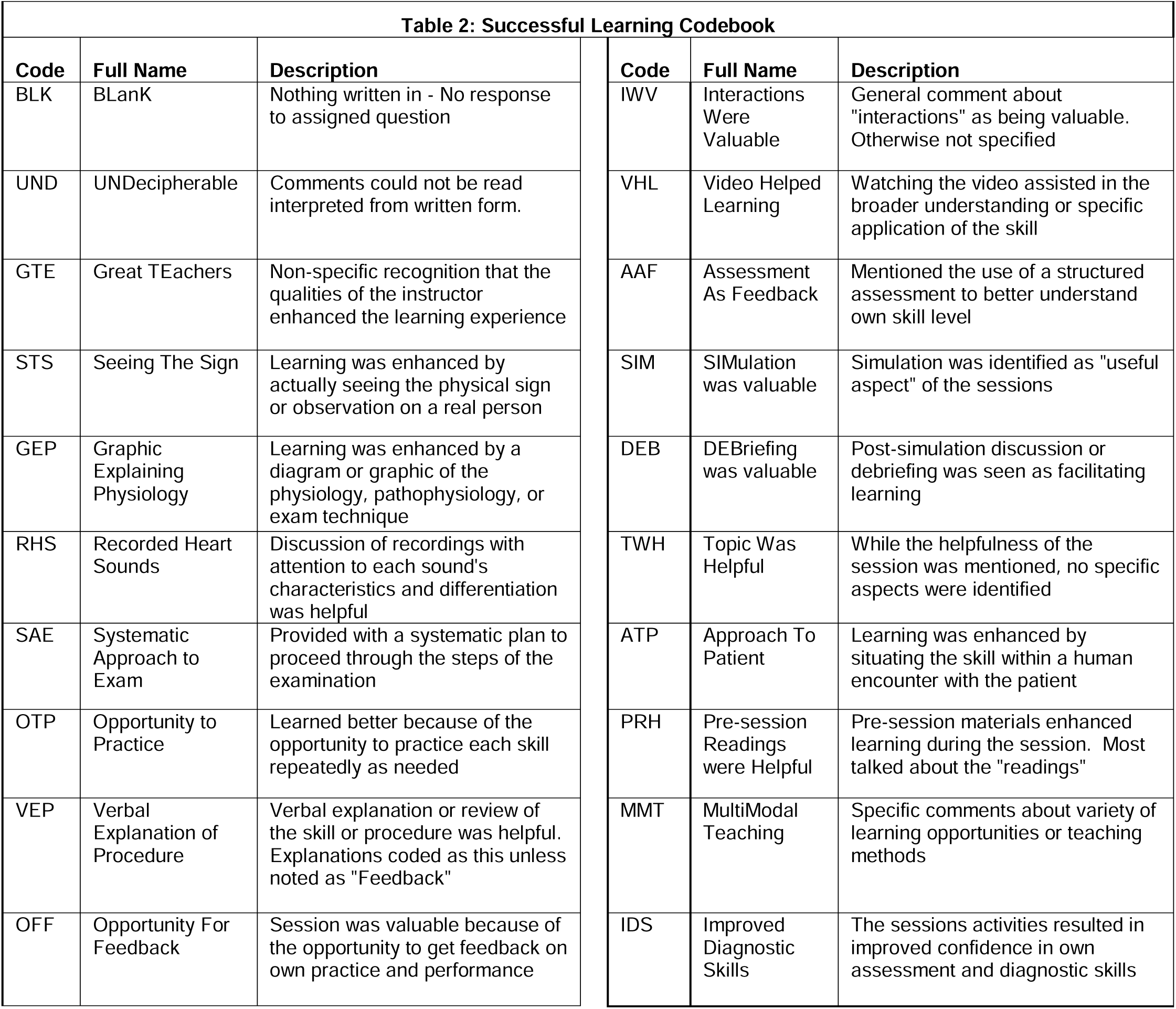
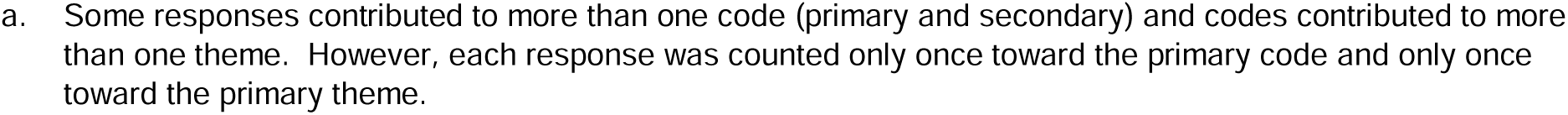
Successful Learning Codebook. Codes were developed using a constant comparison methodology, with the final codebook based on consensus discussions.^a^ See text for details. Third year students on internal medicine rotation, Nov. 2017 - June 2018 Boston University Medical Center, Massachusetts USA

**Table 3.** Opportunities for Improvement Codebook. Codes were developed using a constant comparison methodology, with the final codebook based on consensus discussions.^a^ See text for details. Third year students on internal medicine rotation, Nov. 2017 - June 2018 Boston University Medical Center, Massachusetts USA

| Table 3: Opportunities for Improvement Codebook |  |  |  |  |  |
| --- | --- | --- | --- | --- | --- |
| Code | Full Name | Description | Code | Full Name | Description |
| BLK | BLank | Nothing written in - no response to assigned question | BSC | BedSide Copy was desired | Participant wanted pdf or other portable version of the materials to be able to carry to the bedside for clinical use |
| NIN | No Improvement Needed | Indicated that no improvements were identifiable for the session | ENH | Exercise or activity Not Helpful | A specific activity, practice, or game was not seen as supporting learning and worth the time |
| UND | UNDecipherable | Comments could not be interpreted from written form, or responded as "not sure" | PRB | Pre-Readings/ videos were Burdensome | An excess of pre-class requirements was not helpful. Not enough time to prepare. Too many places to find required items |
| SUS | Poor Teaching/ Scaffolding not sufficient | Instructors did not create effective learning environments or were lacking in some way | ANH | Aids were Not Helpful | Something about mnemonics, graphics, or recordings/videos was not conducive to better understanding of the skill |
| WPC | Wrong Place in Curriculum | Participant indicated that this session would have been more meaningful either earlier or later in the curriculum | Collapsed into ANH, see note above. | Videos were Not Helpful | For whatever reason, the content of the videos was identified as least helpful to learning |
| NEF | Not Enough Feedback | Wanted more opportunity to get feedback of own understanding and skills | NEV | Not Enough Videos | Session could have been improved by additional similar videos |
| DNH | Discussion Not Helpful | Discussion of one or more topics was seen as not contributing to effective learning | SNH | Simulation Not Helpful | Did not find the simulation experience or debriefing helpful, including needing more structure |
| NET | Not Enough Time to think | Wanted more time to understand what was being presented | FPP | Faster Pace Preferred | Session had too much repetition or could have moved faster |
| STD | Steps Too Dense | Too much being taught at one time. More complexity than could be managed as learner | SUB | SURveys were Burdensome | Reported that the surveys were the least helpful for learning |
| NEP | Not Enough Practice | More time practicing skills was desired to gain competency and to meet objectives | CCN | Clarify Course Navigation | Confusing or forgotten instructions |

- a. Some responses contributed to more than one code (primary and secondary) and codes contributed to more than one theme. However, each response was counted only once toward the primary code and only once toward the primary theme.

The emergent codes clustered around various themes in each category. Successful Learning themes centered around Effective Learning Strategies (ELS) and Meaningful Peer Encounters (MPE), skills practice, and educator encounters (see Table 4 for details). Sample comments representing the Successful Learning codes and resultant themes included:

> “The pre-class videos were helpful because they got me thinking what we learned today ahead of time” (code VHL, Participant 1129-08),

**Table 4.**
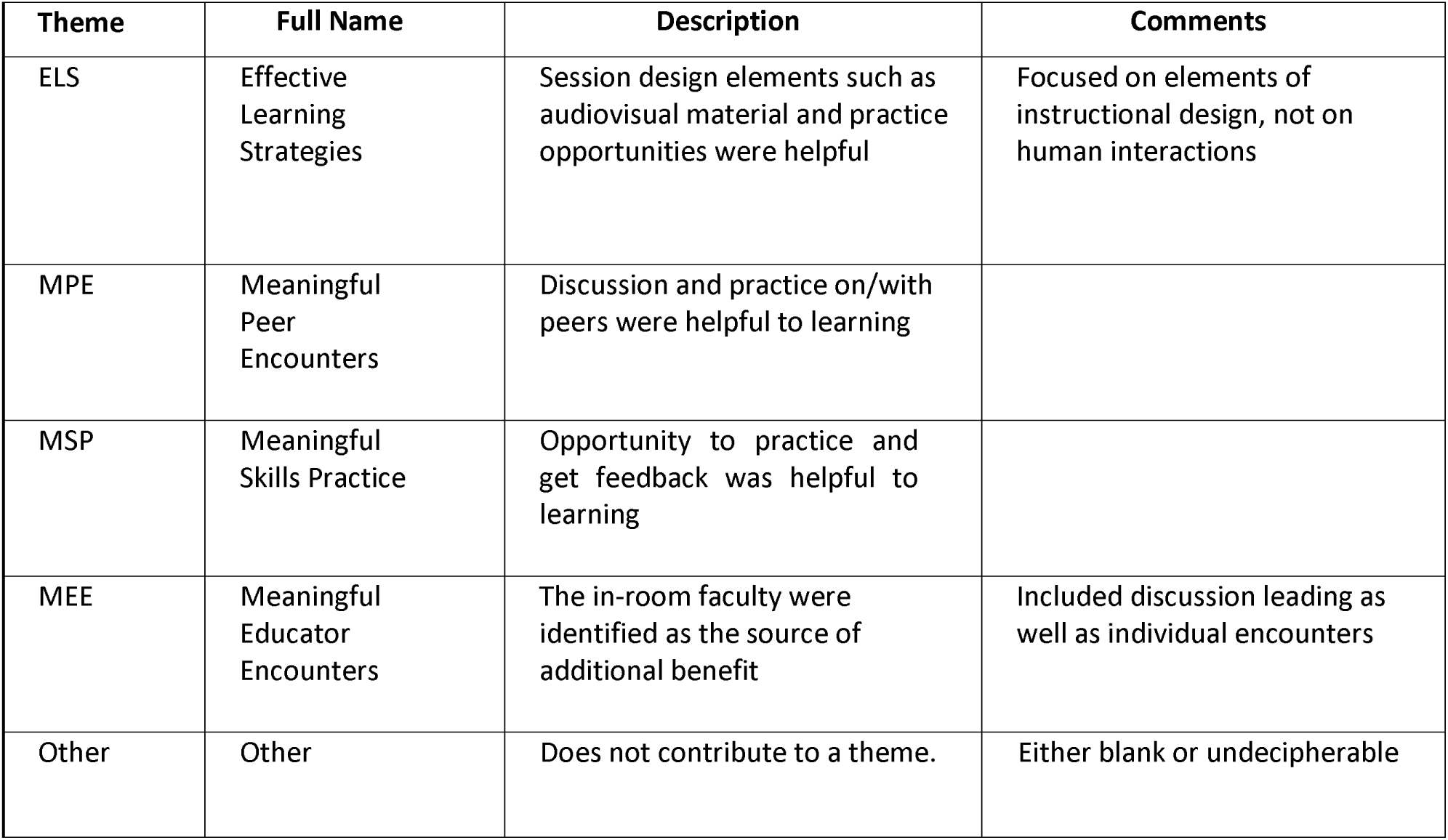
Themes Related to Successful Learning Third year students on internal medicine rotation, Nov. 2017 - June 2018 Boston University Medical Center, Massachusetts USA.

> “I like the standarized [sic] approach to the cardiac exam” (SAE, 120-16),

> “…I prefer to make mistakes in the simulated environment and a little more effective [sic] with active patients (SAE, 1214-10),”

> “Showing examples of approaches to different patients” (ATP, 1206-06), and “Learning how to change the language for different patients” (ATP, 0411-04).

The last three responses also exemplified students’ recognition that holistic care was dependent on effective communication with the patient.

Interactions with others were valued as contributing to learning, as seen here (themes MPE and MEE):

> “The interactive aspect of the in-class activities helped [learning]” (IWV, 1129-11), “In person demonstration…” (DOE, 1129-02), and “Interactive exercises,” (IWV, 1129-10).

Each of these quotes contributes to an overarching concept, the importance of learning beyond the knowledge and skills domains (themes ELS and MSP).

The Opportunities for Improvement themes focused on instructional design improvements, individual learning preferences, instruction related improvements, and general comments regarding how no improvement was needed (Table 5). Sample comments representing the Improvement codes included:

> “Hard to watch videos while in a busy Med service, this would have been more useful in 2nd year when we were learning about physical exam” (WPC, 1129-05) and “… Why should I trust clinical skills likely more relevant of a discussion start of 3rd year” (WPC, 0411-11).

**Table 5.** Themes Related to Opportunities for Improvement Third year students on internal medicine rotation, Nov. 2017 - June 2018 Boston University Medical Center, Massachusetts USA.

| Theme | Full Name | Description | Comments |
| --- | --- | --- | --- |
| IDI | Instructional Design Improvements | Improvements were systems, curriculum, and session design related. | Recommendations could be applied system-wide to the materials |
| ILP | Individual Learning Preferences | Improvements were related to student preferences and needs | Included comments that might be attributable to learning preferences |
| NIN | No Improvement Needed | Answers indicated no adjustments or updates were needed. | Blanks were not included. |
| IRI | Instruction Related Improvements | These were related to the delivery of the experience or session | Attributable to one instructor. Differentiated from systems issues. |
| Other | Other | Does not contribute to a theme. | Either blank or undecipherable |

While the inclusion of the BCA curriculum was seen as integral to the third-year learning, perspectives varied widely on not just the sequence but also the presentation of the concepts within the overall medical school curriculum. This was exemplified by one student’s comment in the Successful Learning category on individual learning preferences and expectations, “The variety of teaching methods used in each session (MMT, 0411-01).” These emergent themes became the basis for our understanding of the learning experiences, as we discuss next.

## Discussion

In this paper, we explore themes that emerged from a cohort of internal medicine clerkship students participating in a curriculum that scaffolded a holistic approach to clinical learning and patient care. Thematic analysis suggested that the learning strategies were effective and that peer encounters, skills practice, and encounters with educators were meaningful. Learners reported that human interaction, and thus learning in the “attitudes” domain, meaningfully contributed to successful learning.

The curriculum explicitly incorporated notions of trust and humility into competency development and underscored the attitudinal, or affective, domain of teaching and learning BCA. Our findings agree with other studies highlighting humility as a vital competency in professional development linked to improved learning, patient-centered care and team collaboration. ^22–24^ The educational approach incorporated such practices as “motivation for cultural humility, …cultural exposure, integrated curricula, case-based learning, …self-reflection, mentorship, and assessments” shown to embed cultural humility into medical education and professional identity formation.^24^ The analysis also revealed opportunities to improve the original instructional design, which were incorporated into the final version of the curriculum.^9^

Taken with an analysis that participants were more confident in their BCA abilities,^9^ the emergent themes suggest the curriculum supports the development of “confident humility,” a trait that the UME-GME Review Committee deems critical to the UME-GME transition.^1^ In a recent integrative review, Matchett and coworkers define humility as the physician’s stance towards the self, others, and the profession. It entails revisiting one’s knowledge and beliefs and is critical for learning, expertise development, and professional growth. Humility is also a linchpin of patient-centered communication and rebuilding trust in the doctor-patient relationship.^8^

The BCA curriculum encourages learners to explore humility and patient-centered communication alongside knowledge and skill acquisition. “Humble physicians recognize that … patients are experts in the meaning of illness within the context of their experiences, values, and beliefs.”^8^ Patient-centered communication, as an attitudinal, meaning-making concept, emphasizes the importance of understanding a patient’s feelings, intentions, assumptions, and perhaps any attributes of the patient that differ from those of the doctor.^10,11^ Promoting such health equity across the UME-GME continuum is a fundamental responsibility of medical schools and GME programs.^25^

All of this suggests that the curriculum may be a useful tool in support of students’ professional identity formation, as elaborated elsewhere,^9^ one that provides several opportunities for “targeted coaching by qualified educators[.]”^26^ Inherent benefits of such coaching would stem from the curriculum’s focus on the learner; potential for improving performance in residency; and, particularly through the “Trust” large-group discussion and simulated clinical encounters activities, explicitly addressing imposter syndrome and creating a safe approach toward addressing areas for growth.^27^

## Limitations

We were unable to explore further the emergent themes from the limited data set, so it is possible that elements of the constructivist—meaning-making—instructional design other than notions of trust were stronger drivers of learners’ experience of the curriculum as “meaningful.”

Given the use of the content analysis approach and inability to examine nuances of the emergent themes, the curriculum’s generalizability to learners in other health professions education programs is limited and may require further study. Further, this dataset was based on responses to similar but not identical survey questions, which may have introduced an unintended bias.

We conceptualized learners’ sense of trust-worthiness as an attitudinal aspect of a competency but acknowledge that it could also be viewed as a construct that is challenging to measure and thus operationalize.^5^ Similarly, while our explicit incorporation of notions of trust in the curriculum aligns with several recommendations for comprehensive improvement of the UME-GME transition,^1^ future concept or outcome analyses are needed to clarify the value of incorporating elements of entrustability beyond their well-established roles in assessment. Finally, even as “humility” is increasingly recognized as “a multifaceted construct [that] may be linked to excellence in clinical medicine,”^23^ we acknowledge the inherent ambiguity, as CBME’s shared language continues to evolve,^2^ of such terms as attitudes, trust, trustworthiness, and entrustability.

### Next Steps

A study of the curriculum’s impact on learners’ BCA-related knowledge, skills, attitudes, and trust-worthiness as a learning construct is ongoing. Other researchers may wish to study the effect of the BCA curriculum on learners’ humility and empathy,^28^ patient-centeredness,^28,29^ or professional identity formation.^30^ More generally, we encourage curriculum developers to use and assess the efficacy of constructivist and patient-centered approaches when developing clinical abilities curricula.^9^

## Conclusion

We developed a BCA curriculum that supported acquisition of professional attitudes as well as traditional knowledge and skills. Learners were urged to hear the patient’s perspective with humility, as a human predicament, and diagnostically, as an element of the clinical history in diagnostic reasoning. A patient’s question, “Why should I trust your clinical skills?” drove the expertise development-related learning outcomes. Content- and thematic analyses of surveys of a cohort of internal medicine clerkship students suggested that the learning strategies were effective and that peer encounters, skills practice, and encounters with educators were meaningful.

The curriculum may be a useful tool in support of students’ professional identity formation. We hope that thoughtful design of curricula that fosters learners’ attitudinal growth will promote its development beyond traditional knowledge and skills, and that this exploratory work will stimulate conversations around expanded roles of notions of trust in medical education.

## Data Availability

All data produced in the present study are available upon reasonable request to the authors.

## Acknowledgements

The study team wishes to thank the many people who were integral to the success of this project, especially the students who learned with us, Sonia Ananthakrishnan, Warren Hershman, Kelly Ho, and David Flynn at Boston University Medical Center (BUMC). Thanks to Tina Grotzer, Flossie Chua, and Bryan Mascio at Harvard Graduate School of Education; Emil Petrusa, formerly of Massachusetts General Hospital and Harvard Medical School; Vlad Valtchinov at Harvard Medical School; Maria Romero at VA Bedford Healthcare System; and Sarah Wood at Harvard Macy Institute. Coauthor Dr. Gail March Cohen, who passed away on 28 March 2024, contributed to project conceptualization, instructional design, and early manuscript review. Coauthors Deborah D. Navedo is currently Founding President, Clinical Perspectives Associates, Boston, MA, USA; Ahmed H. Nahas is a geriatrician currently at Yakima Valley Farm Workers Clinic, Yakima, WA, USA; and Carly M. Eiduson is currently a resident in pediatrics at Children’s Hospital of Philadelphia, Philadelphia, PA, USA. Odindo Opole, Third Joint Productions, produced the video abstract.

## Ethical Approval

The Boston University Medical Center (Boston, Massachusetts, USA) Institutional Review Board (IRB) reviewed the research materials including the participant consent procedures (IRB Number: H-42353, Jan. 10, 2022). The IRB made the determination that the study qualifies as “not human subjects research (NHSR)” based on the definitions of human subject and research under the policies and procedures of the Human Research Protection Program. As an NHSR study, institutional procedures stipulated that individual signed consent was not a requirement, but participating students had to review and accept the following statement prior to participation: “The Boston University IRB has asked us to provide you with the following consent: ‘Participation in the educational research survey is completely voluntary. Data collected will not be linked to your name and will be used for research purposes only.

Participation or non-participation in the survey itself will have NO impact on grades or evaluations.’” Students were explicitly advised that their consent was implied by their participation. Additionally, all quotations used in this analysis were submitted anonymously, transcribed by a non-author, and not identifiable to the authors. Subsequently this work was reviewed by the VA Bedford Healthcare System IRB and determined not to require its oversight.

## Disclaimer

Views and content are those of the authors and do not reflect the official policy or position of VA or any agency of the United States Government.

## Previous Presentations

Meisel JL, Chen D, March G, et al. Building competence and confidence in clinical skills: a systematic, flipped classroom approach to learning bedside cardiac assessment. Poster presented at Northeast Group on Educational Affairs Annual Conference; May 4-6, 2017; Rochester, NY, USA.

Carmona H, Chen D, March G, Bernard SA, Papps N, Meisel JL. New world techniques to teach old world skills: flipped classroom approach to teaching the bedside cardiac assessment. Poster presented at Senior Resident Academic Day, Boston University Internal Medicine Residency Program, Boston Medical Center; May 26, 2017; Boston, MA, USA.

Meisel JL, Chen D, March G, et al. Building competence and confidence in clinical skills: a systematic, flipped classroom approach to learning bedside cardiac assessment. Poster presented at 12th John McCahan Medical Campus Education Day, Boston University School of Medicine; May 31, 2017; Boston, MA, USA.

Meisel JL, Chen D, March G, et al. Building competence: a systematic, flipped classroom approach to learning bedside cardiac assessment. HE795 Scholarly Project presentation to Annual Convocation, Massachusetts General Hospital Institute of Health Professions; June 19, 2017; Boston, MA, USA.

Meisel JL, Chen D, March G, et al. Building competence—an active learning approach. Poster presented at Alliance for Academic Internal Medicine Academic Medicine Week; March 18-21, 2018; San Antonio, TX, USA.

