## Supplementary figures and images for "From Reductionist Skills to Meaningful Learning: Trust and Humility in Bedside Cardiac Assessment" (from another version of this preprint: version 3, 2025-05-07)

### Figure 1. Notions of trust as a central element of both competencies and entrustment decision-making in competency-based medical education

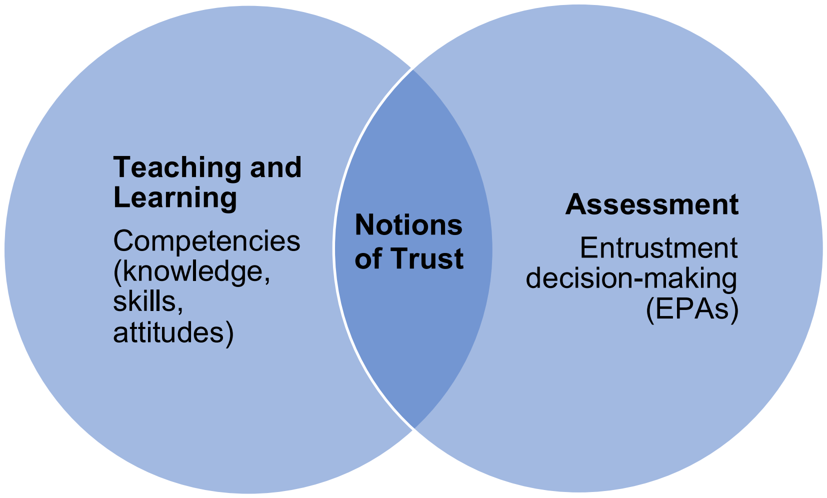
