## Supplementary material for "From Reductionist Skills to Meaningful Learning: Trust and Humility in Bedside Cardiac Assessment": Figure 2. Listen Before You Auscultate bedside cardiac assessment curriculum (from another version of this preprint: version 3, 2025-05-07)

### STEP I: REVIEW LEARNING GOALS

- Describe how to apply a systematic approach to bedside cardiac assessment (BCA) of patients
- Interpret clinical findings related to each step of BCA
- Use the systematic approach to improve your listening and diagnostic reasoning skills
- Correlate cardiac pathophysiology with associated findings on BCA
- Reflect on ways that patient-centered communication is fundamental to bedside cardiac assessment.

### STEP II: COMPLETE LEARNING SETS ASSIGNED VIA EMAIL

Learning Sets Are Labeled "A" through "M." Work through each as follows (90-120 minutes):

1. Read Learning Objectives
2. Activate Prior Knowledge: Question and Answer #1 (Q1 & A1)
3. Watch Short Video
4. Practice and Apply New Knowledge: Question and Answer #2 (Q2 & A2)
5. Read "Linking BCA Findings to Cardiac Pathophysiology: Case" (sets H, I, and J only)

| Before Classroom Session 1, Complete Learning Sets A-F | | | | | | Before Classroom Session 2, Complete Learning Sets G-M | | | | | | |
|---|---|---|---|---|---|---|---|---|---|---|---|---|
| A | B | C | D | E | F | G | H | I | J | K | L | M |
| "Why Should I Trust Your Bedside Cardiac Skills?" | Overview of Bedside Cardiac Assessment (BCA) | Look at the Patient, Feel the Heart | Right Heart Assessment Parts 1 &2 | Left Heart Assessment | Diagnostic Reasoning Tips from Us to You | Listen to the Heart: Left Ventricular Gallops | Systolic Murmurs | Diastolic Murmurs: Aortic Regurgitation | Diastolic Murmurs: Mitral Stenosis | Basic Confirmatory Studies | Heart Sounds Practice & Summary | The Cardiac Cycle (Supplemental) |

### STEP III: PARTICIPATE IN IN-CLASS ACTIVITIES

| Classroom Session 1 | | | | | Classroom Session 2 | | | |
|---|---|---|---|---|---|---|---|---|
| Introduction: BCA and Session 1 (5 min) | Activity 1: Meet Mr. Smith (25 min) | Activity 2: Assess the Right Heart (20 min) | Optional Activity: Question Pool/ Muddiest Point (8 min) | Recap and Looking Ahead (5 min) | Introduction: Session 2 (2 min) | Activity 3: Name that Heart Sound! (16 min) | Activity 4: Simulated Clinical Encounters (38 min) | Recap: Using BCA as a Clinician (2 min) |
